# The Statistical Monitoring by Adaptive RMSTD Tests: an efficient, informative, and customizable method for the complete internal quality control intended for low-frequent sampling of control measures

**DOI:** 10.1101/2020.10.08.20209288

**Authors:** Christian Beier

## Abstract

Two control mechanisms are relevant to perform an internal quality assurance: a permissible limit L_SMC_ applied to single measures of control samples and a retrospective statistical analysis to detect increased imprecision and baseline drifts. A common statistical metric is the root mean square (total) deviation (RMSD/RMSTD). To focus on recent changes under low-frequent sampling conditions, the monitored amount of retrospective data is usually very small. Unfortunately, the calculated RMSTD of a small data set with n<50 samples has a significant statistical uncertainty that needs to be considered in adequate limit definitions. In particular, the minimum reasonable limit L_RMSTD_(n), applied to the RMSTD of a series of n samples, decreases from L_SMC_ (e.g., 2.33*standard_deviation+bias) for n=1 towards L_true_RMSTD_ for n→∞ (long-term statistics). Two mathematical approaches were derived to reliably estimate an optimal function to adjust L_RMSTD_(n) to small sample sizes.

This knowledge led to the development of a new quality-control method: the Statistical Monitoring by Adaptive RMSTD Tests (SMART). SMART requires just one mandatory limit (either L_SMC_ or L_true_RMSTD_) per analyte. By definition of up to 7 possible alert levels, SMART can early recognize and evaluate both the significance of a single outlier and establishing critical trends or shifts in recent SMC data. SMART is intended to efficiently monitor and evaluate small amounts of control data.

## 1 Introduction

Quality assurance, and particularly the internal quality control (IQC), are essential requirements to monitor the reliability of processes in industry and health care. The vast amount of related literature can only be inadequately referred [1-5]. Although, the homepage of Westgard et al. [6] has been established as an informative hot spot with regard to quality assurance in clinical chemistry - demonstrating that it is a versatile and still open topic.

Regulations concerning the IQC in clinical chemistry prescribe consecutive single measurements of at least one control sample (SMC data). Each SMC value is compared to a given maximum permissible limit (L_SMC_) to monitor the measuring uncertainty of an operating device as well as the degradation state of used reagents. A lot of experience already exist to define L_SMC_. The main concepts base on state-of-the-art limits (utilizing performance metrics of the technique), biological variation, and/or partitioned clinical decision ranges. The L_SMC_ delimits an in-control range for SMC results around the target value of the control sample. The target value is predefined either by a reference institution or the manufacturer (labeled control sample). For unlabeled control samples a substitute of the target value can be generated during an evaluation period and possibly additional peer-group data. However, a self-evaluated target value restricts the IQC to a quality monitoring with respect to the state during the evaluation period. Thus, persistent systematic errors of the device, lot, and procedure as well as long-term environmental influences are probably unnoticed.

Another integral part of IQC is a statistical retrospective analysis (RA) of quantitative SMC data at the end of an evaluation period, at the ends of consecutive sampling periods, or by an on-the-fly statistical approach [7-10]. Efficient and widely used statistical metric are the root mean square deviation (RMSD) and the root mean square total deviation (RMSTD), where “total” denotes the optional reference to a known target value instead of the mean. The presented Statistical Monitoring by Adaptive RMSTD Tests (SMART) method is primarily intended for the RMSTD metric. (The RMSTD is sometimes alternatively denoted as the entire analytical measuring uncertainty.) Nevertheless, theoretical findings are provided for both RMSTD and RMSD.

The RA has to handle a trade-off between (i) a sufficient number of SMC samples to reliably verify a statistical in-control condition and (ii) a special focus on most recent data to detect changes in measuring precision as fast as possible. Particularly in clinical chemistry, the RA has to deal with generally low-frequent SMC measures (1-2 SMC per day and control sample) as well as limited reasonable collection periods due to short reagent lifetimes or other frequent system interventions. The presented SMART method has been designed to efficiently evaluate even few amounts of SMC data.

IQC procedures can be combined in multirule concepts [11]. One of the most prominent methods is the “Westgard Sigma Rules” utilizing the Sigma Metric [12]. A detailed and well-structured explanation is given in [13]. The current, improved version is specifically applicable for procedures with different Sigma Metric. The 3s entry rule of the Westgard Sigma Rules (limit range for single SMC values: mean ± 3 times the standard deviation s) is the sole IQC rule for procedures with a high Sigma Metric. However, a high Sigma Metric of six results in a critical systematic error [13] of 4.35 ·s. In other words, the technically achievable precision of the measurement is distinctly higher than necessary compared to the biological variation or medical demands, and one could allow a limited variation in accuracy (bias) and/or a certain level of heteroscedasticity. A potentially allowable tolerance of 1 ·s would be feasible (except analytes that need very precise intra-individual monitoring). Moreover, the applied IQC parameters mean and standard deviation are often self-determined during an evaluation period. This determination is commonly based on a small sample size (n=20-30), which implies a high risk of significantly underestimated parameter values due to statistical uncertainty. As explained in [14], an underestimated standard deviation can actually downgrade the 3s rule to up to a de-facto 2s rule. Both arguments suggest that the 3s entry rule appears rather strict and too general for procedures with a Sigma Metric of six (and even four). Westgard’s multirule concept supports a strictly serial order of decision rules. The SMART method utilizes limit tests in parallel, enabling the possibility to further verify and categorize an out-of-control signal.

Based on a chart of consecutive SMC values, the current standard deviation s_n_ and unsigned bias (mean inaccuracy) δ_n_ can be revealed by 

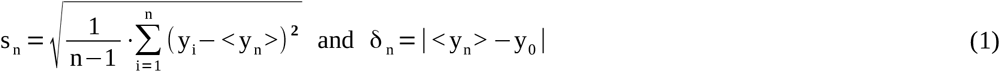

(y_i_: measured SMC, y_0_: target value, <y_n_>: sample mean, n: sample size of SMC chart). Particularly in the case of low-frequent sampling, the parameters s_n_ and δ_n_ are not strictly separable because:

i. The measurement inaccuracy (bias) can vary even within a single chart due to unpreventable or tolerated changes in environmental or operating conditions. These shifts contribute to the obtained amount of the standard deviation.
ii. The average values of arbitrary SMC charts of length n scatter themselves with a dispersion of 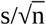. Thus, the amount of the standard deviation affects the obtained average value of a particular chart of limited size - leading to an arbitrary distortion of the real bias of this chart.

It is therefore suitable to solely focus on the entire analytical measuring uncertainty (aka RMSTD) 

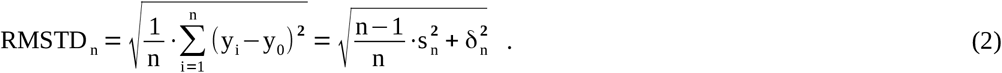

As shown in the right-hand term, the RMSTD can also be expressed as a Pythagorean addition of s_n_ and δ_n_, where the factor (n-1)/n can be neglected for sufficiently large n. However, a separate analysis of standard deviation and bias might nevertheless be useful during fault diagnostics. In addition to the general RMSTD metric, the abbreviation Δ will be used to indicate the “true” RMSTD value that emerges if the statistical uncertainty becomes negligible. Thus, the determination of Δ requires a large data set (see [14] for details).

To indicate out-of-control situations of measuring devices, the IQC usually utilizes at least two maximum permissible deviation limits: one limit intended for each result of an SMC (L_SMC_) and one limit dedicated to the statistical retrospective analysis. This study utilizes the root mean square total deviation; thus, the statistical limits L_RMSTD_ (for arbitrary n) or L_Δ_ (for very large n) are used for this metric. However, since 2008 the German guideline on quality assurance in medical laboratories (Rili-BAEK) [15] only provides one common, maximum permissible limit for both: single SMC and RA. The limit values in the Rili-BAEK, table B1a-c, column 3 are presented as relative total errors Δ^rel^ (in per cent). The absolute value of the solely given, maximum permissible total error is given by 

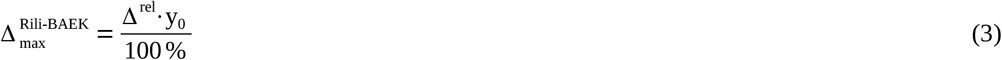

(y_0_: target value of the control sample). The current values Δ_max_ (as prescribed in the Rili-BAEK) are empirically determined and can be treated as approximately equivalent to L_SMC_; however, they are significantly too tolerant if applied as L_Δ_ (see below).

For all analytes without prescribed limits, the self-evaluation of internal laboratory maximum deviation limits is required. These limits are often obtained by following equation, utilizing the bias and the t-fold standard deviation obtained during an evaluation period (t is usually set to 3) 

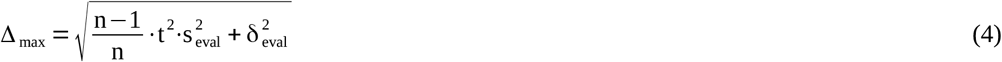

(n: number of SMC values of the evaluation period, s_eval_: standard deviation during evaluation, δ_eval_: bias during evaluation). The factor t with t>1 becomes necessary to receive a reasonably tolerant limit for single SMC measures. However, Eq. (4) with t=3 may provide a still too restrictive limit for SMC (and probably even for RA) in several situations: (i) underestimated s_eval_, δ_eval_ due to poor statistics during evaluation period, (ii) measuring techniques with small imprecision but significant inaccuracy, and/or (iii) a high Sigma Metric of the method (unused room for enhanced tolerance).

As revealed in [16], the implication of the present Rili-BAEK to use the same limit for both the single SMC value and the RA is clearly problematic. In fact, the limits should optimally differ by a factor of about two, where L_SMC_>L_Δ_ (see [16] and Chapter 2.1). This finding and the given statements to handle outliers require a complete revision of the currently prescribed RA of the Rili-BAEK [15]. Seeking a more powerful approach, the SMART method has been newly developed and discussed here in detail. SMART is an RMSTD-based method facilitating evaluation of single SMC results as well as on-the-fly statistical monitoring of recent SMC data for each analyte. It is sufficient to apply SMART with only one prescribed maximum permissible limit. Thus, two alternate versions exist, utilizing either L_SMC_ or L_Δ_ as the prescribed limit.

## 2 Materials and methods

### 2.1 The relation between optimal limits for SMC and RA

All limits defined in this article are unsigned values specifying a range around the target value y_0_±L_given_. The presented SMART method is intended to provide a common approach for the entire IQC, considering only one prescribed maximum permissible limit. It is based on the finding that an approximately constant general relation exists between the optimal maximum permissible limits for one SMC value (L^max^ SMC) and for the true RMSTD of a (hypothetically) very large retrospective set of SMC data (L^max^ Δ) [16]. Hence, the parameter λ is defined as the general relation factor 

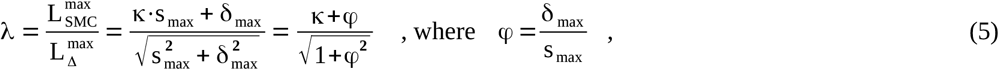

(s_max_: maximum permissible standard deviation; δ_max_: maximum permissible bias; κ: constant expansion factor according to the desired confidence level). The sign Δ (true RMSTD) emphasizes that - assuming a very large considered sample size - there is no remaining statistical uncertainty in the determination of the RMSTD, standard deviation, and bias. In such a case of a very large data set, the confidence interval (CI) of Δ becomes negligible, and the limit L_Δ_ or L^max^_Δ_ can be reduced to be almost equal to the expected exact RMSTD as given by Δ^2^=s^2^+δ^2^.

The factor λ only depends on the ratio between δ_max_ and s_max_. This is shown in Eq. (5) by substitution with the ratio φ=δ_max_/s_max_. Fig. 1 illustrates the mathematical relation between λ and φ for 2 common κ. The factor κ in Eq. (5) can be chosen according to a one-sided CI, due to a generally distinct maximum permissible bias, which is often similar or higher than s_max._. A value between 1.96 (97.5% confidence) and 2.33 (99%) is convenient to avoid regular false-positive outliers. For φ-ratios significantly below 1, a two-sided CI is more realistic, which would change the CI level for κ=2.33 to 98%.

**Figure 1:**
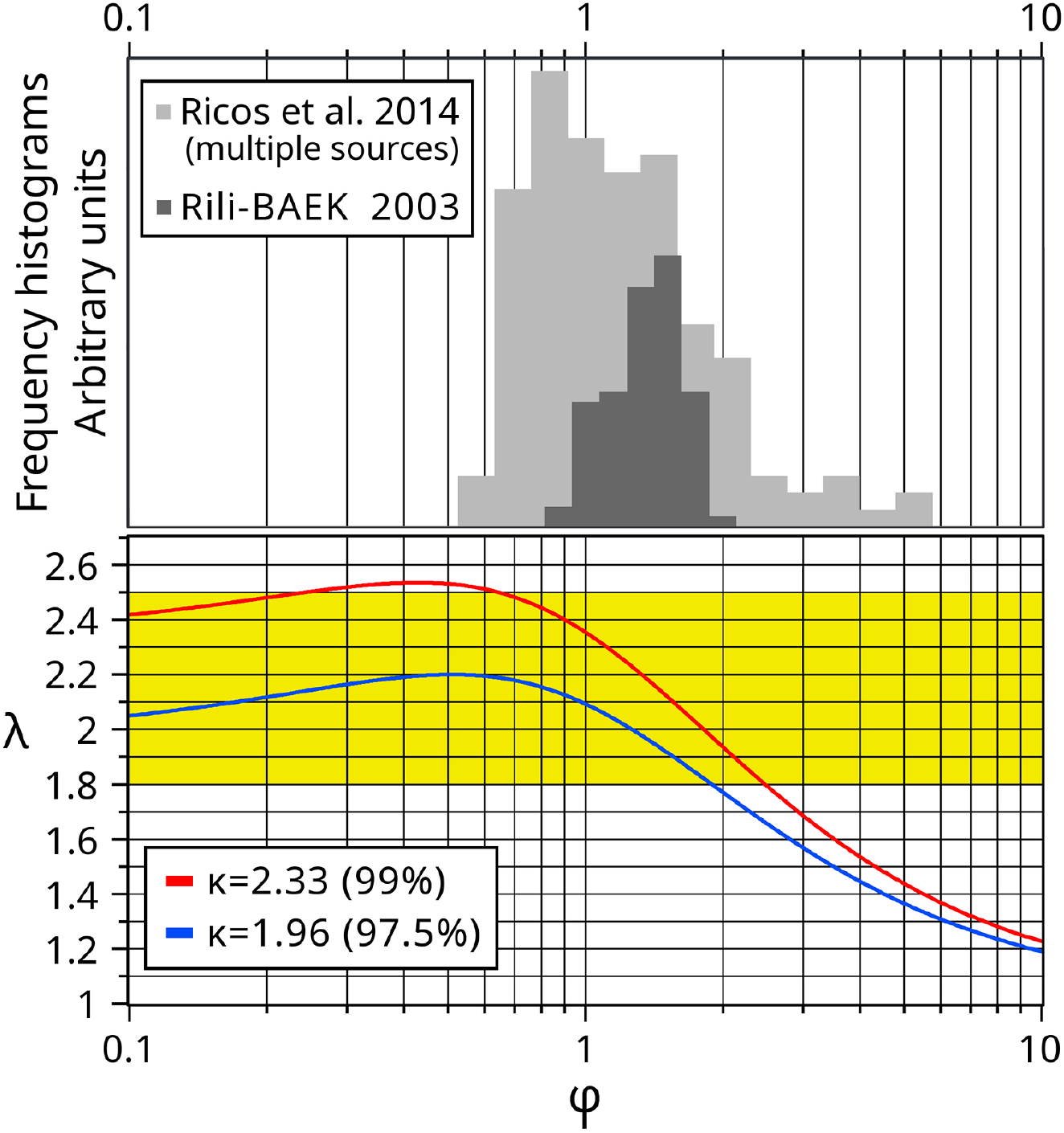
Common relations between maximum permissible bias δ_max_ and standard deviation s_max_ in clinical chemistry are utilized to approximate a general value of λ. (A) Frequency histograms of ratios φ=δ_max_/s_max_ of medical analytes taken from Ricos et al. 2014 (light gray, 150 entries, mean=1.1) and the German Rili-BAEK 2003 (dark gray, 90 entries, mean=1.34) (see Chapter 2.1). Histogram sampling and statistics were done in log_10_ space. (B) Functional relations between λ and φ for 2 different κ according to Eq. (5). Related one-sided CI’s are given in parenthesis. The ratios φ are presented on a log_10_ scale.

To get a profound idea about the distribution of φ over almost the entire spectrum of analytes in clinical chemistry, Fig. 1 shows frequency histograms of φ-ratios based on data of Rili-BAEK 2003 (column 5/6) [17] and the database of desirable limits by Ricos et al. (version 2014) [18,19]. In the latter case, the considered data are limited to entries, which are referenced by at least two independent sources according to column 3 [18]. This limitation is based on the assumption that the finally considered entries represent validated and established techniques to a greater extent. This “multi-source” data set consists of 150 remaining entries. Except slightly less populated margin areas, the histogram of the multi-source data is similar to the full-data histogram (compared in [16]).

The Rili-BAEK data of 2003 (last official declaration of separate δ_max_ and s_max_ permissible limits) represent defined state-of-the-art limits for 90 common analytes (however, at a technological state 15 years ago). Although the absolute limit values are outdated, the φ-ratios are assumed to stay approximately valid, due to limited progress in a disproportionate bias prevention since 2003. Using Eq. (5) the Rili-BAEK data reveal λ values in the range 2.19±0.25 (for κ=2.33) and 2.0±0.2 (for κ=1.96).

The desirable maximum bias and standard deviation entries by Ricos et al. (updated 2014) [18,19] were derived from biological variations (see [20,21]). The 95% confidence intervals of λ based on the multi-source dataset are 2.23±0.5 (for κ=2.33) and 2.0±0.4 (for κ=1.96). The approach used by Ricos et al. that utilizes biological variations [20,21] is popular. However, the applied formulas are not flexible enough to handle the entire spectrum of analytes, ranging from analytes with extremely small ratios of intra-individual vs. inter-individual biological variation to analytes with dominant intra-individual variation (e.g., due to circadian rhythms or prandial state). The analyte-specific desirable maximum limits for bias and standard deviation (based on biological variation) are usually significantly more divergent, compared to curated state-of-the-art limits (see Fig. 1). In particular, the desirable limit for the bias is often too liberal. Hence, statistics were redone under the restriction that the bias is not allowed to be more than two times higher than the standard deviation (accepted ratio φ≤2). This restriction excludes 17 of 150 entries (11.3%). The full ranges of remaining λ’s are then 2.30±0.34 (for κ=2.33) and 2.05±0.26 (for κ=1.96). Please mention that a φ-ratio above 2 can also be circumvented by accepting a more tolerant standard deviation.

Particularly with regard to the applied state-of-the-art permissible limits, one can finally conclude that φ values of current measuring procedures are narrow enough to estimate a general λ value for almost every analyte in clinical chemistry. Taking a rather restrictive significance level that only allows about 1-2% false-positive outliers, the general λ value is within 

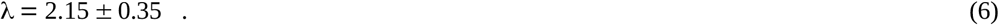

This sufficiently well defined relation between L_SMC_ and L_Δ_ facilitates the explicit definition of only one limit per analyte (either for SMC or long-term RMSTD) for the entire internal quality control. Because of the special shape of λ(φ), the given range in Eq. (6) will remain valid even if future progress in clinical chemistry succeeds to significantly reduce measurement biases. If only the standard deviation needs to be monitored, λ would be equal to the κ value (two-sided CI).

### 2.2 Basics of the Statistical Monitoring by Adaptive RMSTD Tests

The SMART method provides a single formula for both the quality control of single SMC results as well as an on-the-fly statistical monitoring of recent SMC data. Unfortunately, the calculated RMSTD of small SMC series with n samples has an increased uncertainty (due to poor statistics) that needs to be considered in adequate limit definitions. In particular, optimal limits depend on n and exponentially decrease from L_SMC_ (at n=1) to L_Δ_ (for n→∞). Under consideration of the given factor λ (see Chapter 2.1), it is sufficient to explicitly prescribe just one of the two limits.

Provided that a reliable method is given to define n-dependent intermediate limits, the general SMART formula 

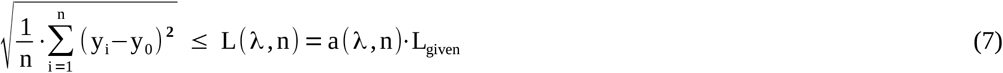

utilizes RMSTD tests against such variable limits L(λ,n), which depend on λ (a constant) and the varying sample sizes n of the considered most recent SMC values. Again, the applied limits must range from the higher L_SMC_ (n=1) to the significantly lower L_Δ_ (n>>1). Thus, a certain limit L(n), used for a series of n samples, has to become increasingly less tolerant at larger sample series. Intermediate limits L(λ,n) are expressed by the transition or adaptation function a(λ,n) that modifies one predefined limit L_given_ (see next chapters). The curvature of a(λ,n) with regard to n has been deeply investigated by two different mathematical approaches as discussed in Chapter 2.3.

After each new SMC measure, Eq. (7) is applied several times considering increasing numbers of previous SMC measures. Thus, the SMART approach performs multiple limit controls, considering the new SMC value (n=1) and up to n_max_ latest SMC values of the same control sample. The recommended maximum size of the entire retrospective window (i.e., the maximum number of recognized, most recent SMC values) is n_max_=15.

Particularly, for each new SMC value, the SMART formula is applied up to 8 times for chart lengths of, e.g., n={1,3,5,7,9,11,13,15} including the retrospectively accumulated SMC values {y_i_}, {y_i_,y_i-1_,y_i-2_}, {y_i_,y_i-1_,y_i-2_,y_i-3_,y_i-4_ }, …, {y_i_,y_i-1_,…,y_i-n-1_}. Hence, up to 8 potential violations of the n-dependent limit levels might be revealed according to Eq. (7). The final evaluation of all 8 limit tests is done by the definitions of alert levels given in Table 1. Alert levels and an efficient use of SMART during the initial phase (available data below n_max_) are discussed in Chapter 3.3.

**Table 1:** Alert levels of SMART

| Level | Indication | Description |
| --- | --- | --- |
| 0 | In control | No violation of limits |
| 1 | Trend warning | Up to two limit violations except at n=1 |
| 2 | Suspicious measure | Outlier at n=1 but no other limit violation |
| 3 | Problematic measure | Outlier at n=1 and one further violation |
| 4 | Statistically out-of-control | More than two limit violations except at n=1 |
| 5 | Fully out-of-control | Outlier at n=1 and at least two further violations |
| 6 | Deprecate lot/device | Exceeding a given maximum permissible number of level-4 or level-5 events within last, e.g., 100 days of operation |

Other variants of SMART settings (the “SMART plan”) are possible. The limit tests do not need to apply equidistant expansions in sample size. Nevertheless, it is strongly recommended to keep the first two tests at n=1 and n=3. To provide an efficient evaluation of SMART results, the number of applied limit tests per SMART run should be at least 6. Another SMART plan of practical interest might be a monitoring window of 19 most recent SMC values and 7 limit tests. These tests could accordingly be done via successive increase of the analyzed data by additional 2-4 retrospective values n={1,3,6,9,12,15,19}.

### 2.3 The curvature of the adaptation function

The considered data set of an RA is often very small on a statistical point of view (n<30). It results from a usually small retrospective time window (to ensure sufficient topicality) and/or a small available number of recent SMC results due to low-frequent sampling. This has consequences for the applied maximum permissible limits of statistical metrics, which are usually defined for “sufficiently long” charts of SMC data (i.e., with limited or no consideration of statistical uncertainties). The minimum amount of collected SMC data to reliably apply such limits is commonly distinctly underestimated (see Chapter 3.1). Regarding SMART we thus need to know, how RMSTD limit levels L(n) have to be increased when they are applied to smaller and smaller sample sizes (due to the increased statistical uncertainty). It is assumed that the limits merge with the limit L_SMC_ at n=1. The functional dependency of L(n)=a(n)·L(n=∞) on n is estimated by two different mathematical approaches outlined in the following chapters. These approaches are fully explained and derived in [14].

Both approaches utilize a rather tolerant CI of 95%. The author recommends limits based on a relatively narrow CI like 95% in combination with a constant small addition to all evaluated parameters. The addition might be in the order of the difference in maximum amplitudes (at n=2) between the CI-99% and CI-95% functions. Please also note that the final scale (i.e. range of amplitudes) of SMART limits is actually only based on λ.

#### 2.3.1 The maximum error propagation of statistical uncertainties

The statistical limit L_RMSTD_(n) must be equal or higher than the RMSTD of any possible in-control SMC data set with n considered values. The maximum expected RMSTD value of a limited data set can be approximated by the upper limit of the confidence interval (CI_Δ_^up^) of the RMSTD metric. In our case the CI_Δ_^up^ of the closely in-control true RMSTD value. The maximum in-control true RMSTD value is predefined but only applicable to a sufficiently long SMC chart. It is therefore necessary to estimate the CI_Δ_^up^ for small sample sizes n, which will finally provide inference with regard to the curvature of the adaptation function a(λ,n) of SMART.

In [14], a rather straightforward approach is presented to determine the true RMSTD value and to approximate the CI_Δ_^up^ of the RMSTD metric. According to [14] the final function of CI_Δ_^up^ with regard to the variable n results in 

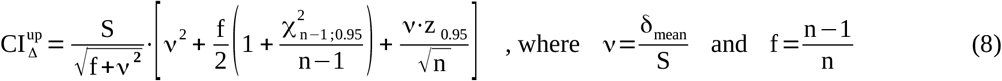

(n: number of considered SMC values, δ_mean_: mean bias (unsigned difference between the overall mean and the reference value), S: overall standard deviation, χ^2^ and z: quantiles of the chi-squared and normal distributions). The parameters δ_mean_ and S may be evaluated by a big-data multi-chart analysis of the measuring system as described in [14].

It is important to mention that the ratio ν=δ_mean_/S is not directly comparable to φ, the ratio of the maximum permissible single-chart values δ_max_ and s_max_ given in Eq. (5). The individual biases of single SMC charts vary according to device-specific, operational, and environmental conditions with a scatter parameter S_δ_. The procedure, presented in [14], includes SMC data from several SMC charts. In the overall statistics, the variability between biases of individual charts is thus part of the overall standard deviation term S. This implicitly included variation of biases can be treated as normal-distributed. Pure short-term dispersion (imprecision) and the variation of biases are further sufficiently independent; thus, the overall variance is the sum of both variances S^2^=S^2^_imp_ +S^2^_δ_. The remaining mean bias δ_mean_ is just the difference between the overall mean value and the target value. If the target value is a pre-evaluated nominal value, δ_mean_ is expected to be very small in relation to S. However, the amount of δ_mean_ might be still significant, if the target value was revealed by a more precise reference method. In general, the ratio ν=δ_mean_/S is expected to be clearly non-zero but distinctly smaller than the ratio φ (defined in Chapter 2.1) 

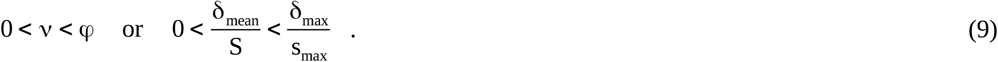

With regard to common relations between δ_max_ and s_max_ as presented in Fig. 1, a proper value for v is approx. 0.6 with an assumed maximum uncertainty of about ±0.5. Further, the decision to apply a one- or two-sided 95% CI for the parameter z_95%_ also depends on v. If the amount of δ_mean_ is almost as high as S (or even higher), a one-sided CI is more suitable than a two-sided CI. If one apply a smooth transition between two-sided (ν=0, z=1.96) and one-sided (ν=1, z=1.645) statistics, the dependency of CI_Δ_^up^ on such ν-z combinations is relatively small, with a broad central maximum at about ν=0.3-0.5 [14]. The combination of ν=0.6 and z_95%_=1.7 (or z_99%_=2.4) is considered as a feasible representative choice for all relevant ν-z combinations.

As already mentioned, a limit function for RMSTD values must be equal (or higher) than the upper CI limit of Δ. To focus on the curvature of CI_Δ_^up^ that only depends on the ratio between δ mean and S, the CI_Δ_^up^ function is normalized by its value at infinite n. At n→∞ all statistical uncertainties can be neglected and CI_Δ_^up^ becomes equal to the true RMSTD: 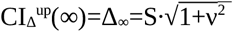. Thus, the relative limit function is finally 

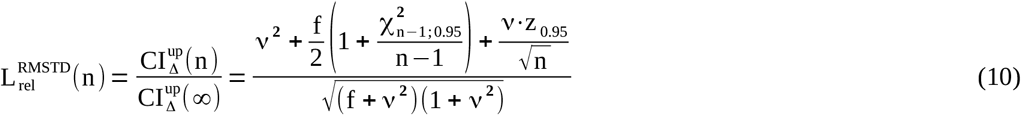

(n: sample size; f=(n-1)/n; ν=δ_mean_/S; χ^2^ and z: 95%-quantiles of the chi-squared and normal distributions).

The given mathematical approach is intended to estimate the statistical uncertainty of an RMSTD of small series. However, the presented model becomes itself distinctly uncertain for any n below about 6, due to (i) the pragmatic concept of the degrees of freedom (esp. correction factor f) and (ii) to the fact that the underlying Taylor-series development is limited to the first degree, thus less intended for large uncertainties. Therefore, an alternative mathematical approach is introduced in the next chapter to further prove the revealed curvature.

#### 2.3.2 The Multidimensional Confidence Interval

To further validate the curvature of the adjustment of SMART limits with respect to small sample sizes, a second approach has been derived. This alternative theory extends the CI of a single measure to a common CI of a series of n measures, while keeping the overall confidence level constant (at, e.g., 95%). It will be denoted as the Multi-Dimensional Confidence Interval (MDCI). The approach requires two general assumptions: (i) the total measuring uncertainty is purely normal-distributed around a constant mean and (ii) all consecutively measured results are uncorrelated (i.e., independent measures). The MDCI finally leads to the upper limit function L° of the root mean square metric. Due to (i) the MDCI theory neglects the amount and uncertainty of the mean bias; thus, it only represents an RMSD-like metric.

The MDCI theory originates from the calculation of the two-sided CI of a normal-distributed and standardized variable by integration of the Gaussian probability density (GPD) function. Applying the known statistical parameters S (overall standard deviation) and δ_mean_ (mean bias), revealed by a sufficiently large evaluation set of SMC values (see [14]), each measured SMC value can directly be standardized without loss of information. The MDCI theory is therefore simplified to realizations of the standard Gaussian distribution (where RMSD(n→∞)=1).

The combined probability of n independent events (occuring sequentially or in-concert/multivariate) is the product of the single-event probabilities P(1,2,…,n)=p_1_·p_2_·…·p_n_. If the single p-values are extended to integrals over one-dimensional GPD distributions, the combined probability P leads to an n-dimensional GPD distribution. Thus, the total probability of a particular sequence of n standardized control measures can be expressed by this n-dimensional GPD distribution, where each dimension represents one control measure of the SMC sample series. Any n-dimensional GPD distribution is rotationally invariant around zero and can therefore be expressed as a radial function. Here, we are only interested in isolevel/equipotential spherical hypersurfaces within the spanned space. This is for example the spherical partitioning surface of a certain common confidence limit (z) represented by a constant radius. To obtain the (two-sided) 95% confidence limit z_95_(n), the normalized radial density function has to be integrated from 0 to z_95_(n) 

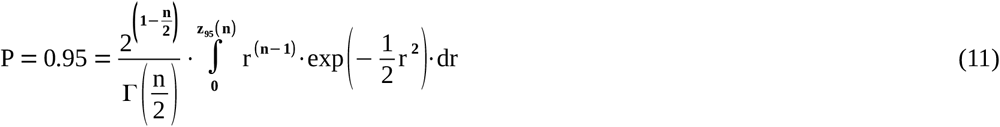

(n: sample size or dimension; r: radius of the spherical hypersurface; Γ: Gamma function). Thus, the radial integration limit z_95_(n) covers 95% of the hypervolume spanned by the multidimensional GPD distribution. This slightly more advanced theory has been fully explained in [14], including a complete derivation and validation of Eq. (11).

Obviously, z_95_ is 1.96 at n=1. For higher n, the limit z_95_(n) increases according to Eq. (11) and slowly converges towards the square root function of n. According to [14], the limit z_95_(n) represents the Euclidean distance r_CI_ of a series of single confidence limits ensuring a constant overall significance level of 5%. The limit can therefore be utilized to obtain the maximum acceptable RMSD (with 95% confidence) of a sequence of n SMC measures. To obtain such a function of adequate upper RMSD limits L°(n) of n standardized values, z_95_(n) can directly be implemented into Eq. (2) (considering y_0_=0) leading to 

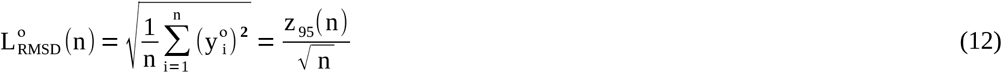

(° denotes the RMSD limit of an N(0,1)-distributed sample; y_i_°: standardized SMC values). L°(n) goes from 1.96 (n=1) to 1 (for n→∞) as drawn in Fig. 2A. The curvature of L°(n) is assumed to be directly comparable (except a different scale) to the adaptation function a(λ,n) of SMART as defined in Eqs. (7) and (13). The radial limit z_p_(n) is also equivalent to the square root of χ^2^ (n). Relevant metrics (Γ(n/2) and pairs of z(n), L°(n) for the 3 two-sided CI’s 95%, 97,5%, and 99%) for all n≤40 are provided in [14].

**Figure 2:**
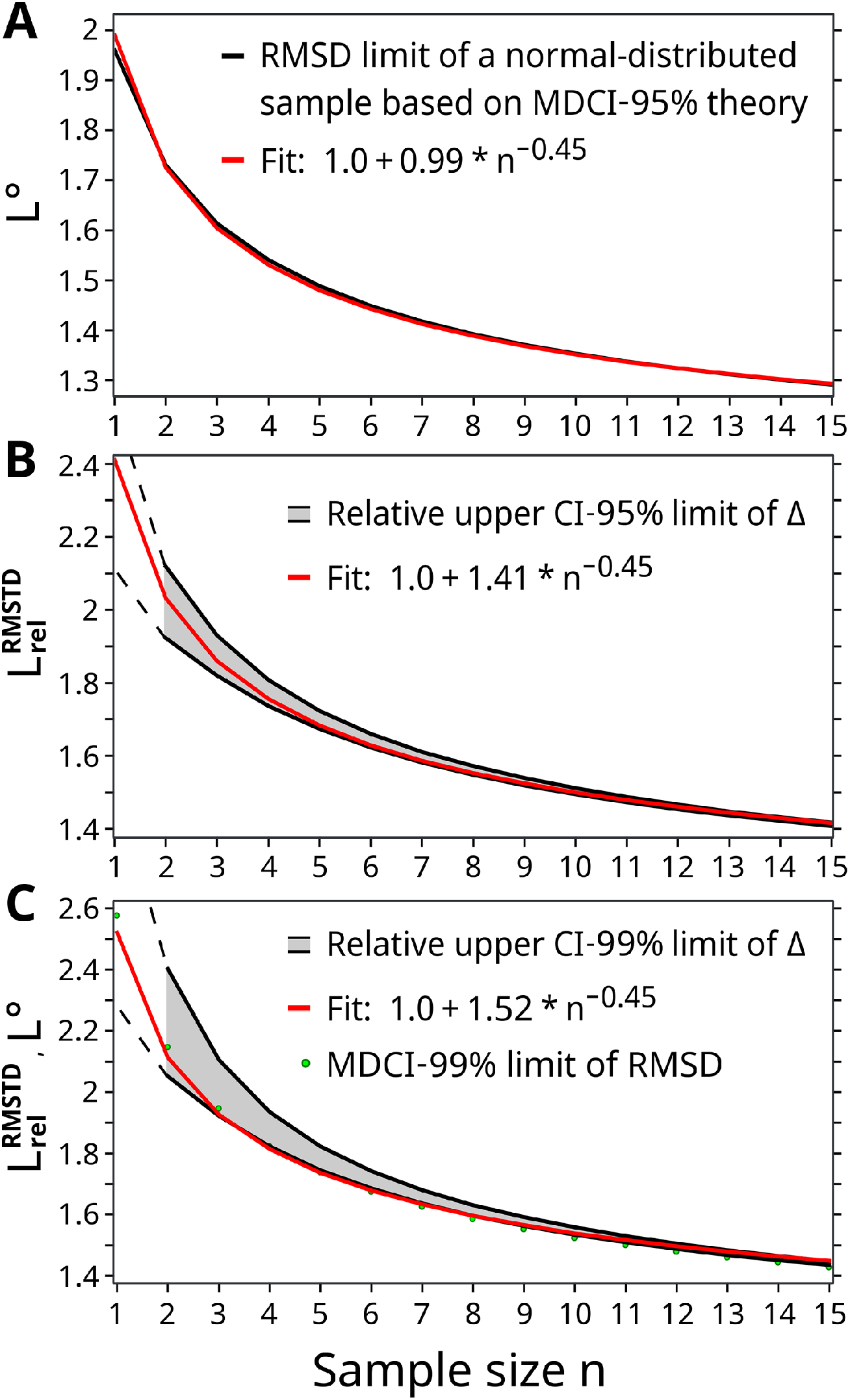
The graphs show the relative increase of the statistical uncertainty to determine a reliable value of RMSTD/RMSD based on small sample sizes n. All presented functions converge to 1 at n→∞. (A) The graph is related to the MDCI theory at a confidence interval of 95% (black line). (B,C) The black functions represent the statistical uncertainty of the RMSTD metric given by the CI_Δ_^up^ approach with ν=0.6. The CI_Δ_^up^ approach becomes itself inherently uncertain at very small sample sizes n<6, indicated by gray corridors and explained in [14] and Chapter 3.1 Further, a value for n=1 is not defined by the CI_Δ_^up^ approach. Thus, the dashed lines mark extrapolations by multi-parametric fits of the boundaries of the corridors. The corridors in (B) and (C) differ in the related confidence interval and the type of error propagation: (B) 95%, maximum error propagation; (C) 99%, Gaussian error propagation (see Chapter 3.1). Graph (C) also includes the MDCI function at CI-99% indicated by green dots. A best-matching function with same curvature (n**^-0.45^**) is aligned to all graphs (red lines). This type of function is used in the SMART method.

### 2.4 SMART utilizing the limit of the expected long-term RMSTD

The “true” RMSTD, which emerges by a drastically extended sample size (n>>100), can be revealed rather precisely by Eq. (2) and the simple evaluation method described in [14]. This method evaluates the necessary parameters s and δ (together with a post-processing of data) with high precision by a Gaussian fit of the overall histogram of multiple long SMC charts. Thus, the maximum permissible statistical limit L_Δ_ for a (hypothetically) very long SMC chart can be set rather strict. Only a small additional amount (i.e., permissible bonus) is sufficient to provide a robust definition of the general L_Δ_ limit per analyte. Such a moderate bonus facilitates some tolerance for heteroscedasticity or baseline drifts. This SMART variant, which adapts the given L_Δ_ to small SMC charts, is recommended due to the high reliability of large-scale statistics used to determine L_Δ_.

As further discussed in Chapter 3.1, the given mathematical analyses to approximate the expected curvature of a(λ,n) lead to the following final equation of this SMART variant that refers to L_Δ_ 

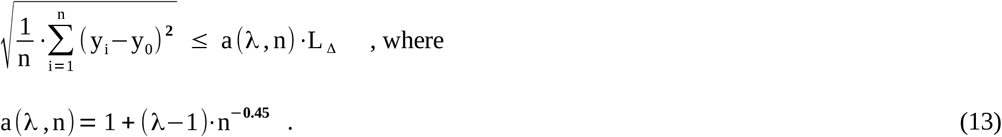

The adaptation function a(λ,n) modifies the long-term limit L_Δ_ with regard to smaller and smaller sample sizes n. It implicitly goes towards the higher limit L_SMC_ at n=1. A proper general value for the constant λ can be obtained from Eq. (6). To be valid for (almost) all analytes in clinical chemistry, a λ value near the upper limit of the given uncertainty range of λ is recommended (i.e., λ closely below 2.5).

### 2.5 SMART utilizing the limit dedicated for single measures (L_SMC_)

The SMART method can alternatively be provided with sole regard to the maximum permissible limit L_SMC_ intended for single measurements of control samples. Such limits are often the only statutorily prescribed information in guidelines. For example, the limits given in the German guideline (Rili-BAEK, table B1a-c column 3, in relative values [15]) can be considered as sole L_SMC_ data [16]. In contrast to the determination of L_Δ_, the definition of L_SMC_ is usually done empirically at a certain significance level and/or related to a fraction of the biological variation of the analyte [20,21]. The determination of L_SMC_ might also base on Eq. (4), where s_eval_ and δ_eval_ could be revealed by a Gaussian fit of the overall histogram of evaluation data. However, Eq. (4) yields too strict limits in special cases (small imprecision but significant inaccuracy). Depending on the technical precision of the examined measuring process in relation to the medical needs (quantified by, e.g., the Sigma Metric [13]), a small addition could be permitted to its L_SMC_.

Running SMART, the applied limits decrease with higher considered sample sizes n from L_SMC_ (n=1) towards L_Δ_ (n>>1). Here, L_Δ_ is implicitly given by the constant λ according to Eq. (6). The preliminary suggestion in [16] to apply a natural exponential function for a(n) was based on the assumption that the uncertainty of the RMSTD at a sample size n=15 has already reached a value near the long-term level with just a small remaining slope. However, one of the main new conclusions, presented in Chapter 3.1, reveals that the remaining offset and slope are still very significant at n=15 compared to n→∞. The curvature of a natural exponential function appears now to be distinctly too steep in the range n=1-15 with the clear risk to cause false-positive limit overruns at higher sample sizes.

The newly revealed functional dependency of a(λ,n) is strongly justified by the two presented mathematical approaches (Chapter 2.3). The final equation of SMART utilizing L_SMC_ and dedicated to the range n=1-15 is given by

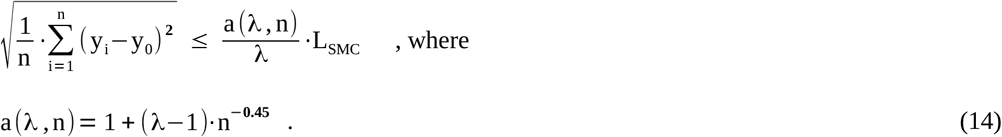

The choice of λ depends on how tolerant the given limit L_SMC_ has been defined. Assuming a rather restrictive value of L_SMC_, λ should be chosen near its lower boundary given in Eq. (6) to prevent false-positive results for (almost) all kinds of analytes (i.e., λ=1.8).

## 3 Results and discussion

### 3.1 The uncertainty of the RMSTD due to small sample sizes

In Chapter 2.3 and [14], two mathematical approaches were derived to quantify the increased statistical uncertainty of RMSTD results at relatively small sample sizes. The quantification of this uncertainty is important to optimally adapt the applied IQC limits for RMSTD results of short SMC charts (especially for n<<50). The derived approaches are denoted as CI_Δ_^up^ (Chapter 2.3.1) and MDCI (Chapter 2.3.2). The first approach refers to the entire RMSTD (inclusive bias); however, it lacks precision below about n<6. The second approach (MDCI) represents a more fundamental theory, but it neglects a systematic bias with respect to a target value (just RMSD). In the comparable case of zero bias (ν=0 in Eq. (10)) and a CI of 95%, the final functions of both approaches are in very good agreement as demonstrated in [14].

The most important conclusion regards to the functions of optimal RMSD/RMSTD limits depending on the considered sample size. Figure 2 summarizes the revealed curves of both mathematical approaches, which are (2A,2C) the standardized RMSD limit L°(n) of MDCI utilizing Eq. (12) and (2B,2C) the relative RMSTD limit (L_rel_^RMSTD^) of CI_Δ_^up^ utilizing Eq. (10). The graphs 2A and 2B represent a relatively moderate increase of the limits by using a CI of 95%, which is recommended in combination with a small constant addition to the prescribed value of the IQC limit L_SMC_ or L_Δ_. The limit functions in 2C provide more tolerance due to a CI of 99%. Please note that - along with the different CI’s - the type of error propagation of the CI_Δ_^up^ approach has also been changed from maximum error propagation (CI-95%) to Gaussian error propagation (CI-99%). The alternative equation of L_rel_^RMSTD^ based on Gaussian error propagation can be found in the appendix of [14]. In this study, L_rel_^RMSTD^ has always been applied with the reference set ν=0.6 and z_95%_=1.7 / z_99%_=2.4, suitable for common analytes in clinical chemistry (see Chapter 2.3.1 and [14]). In Fig. 2B and 2C the intrinsic uncertainty of the CI_Δ_^up^ approach with regard to the concept of the degrees of freedom of the χ^2^-term (discussed in [14]) is visualized by an upper and lower boundary (gray corridor) flanking the expected shape of L_rel_^RMSTD^. The upper boundary utilizes the given Eq. (10), where the lower boundary represents Eq. (10) with a substitution of the term χ_(n-1)_^2^/(n-1) by χ_n_^2^/n.

The curves in Fig. 2 are fitted separately with the same fit function of the type F(n)=1+C·n**^-0.45^** with a best-matching scaling parameter C. All fits in Fig. 2 (red lines) clearly show that the curvatures of the two different mathematical approaches are very similar. Only the amplitudes differ by up to 20% (at CI-95%), which can be dedicated to different basic assumptions. SMART uses an alternative (independent) concept for scaling under consideration of the parameter λ (see Chapter 2.1). Thus, only the curvatures of the limit functions are relevant in this study. Nevertheless, the recommended range for λ (1.8-2.5) agrees well with the (partially extrapolated) amplitudes at n=1 and CI-95% of both mathematical approaches. Fig. 2 confirms that the type of the fit function F(n)=1+C·n**^−0.45^** is very convincing. The exponent (−0.45) can be defined rather precisely. Curvatures with exponents outside the range (−0.46,-0.43) are clearly less matching. An additional feature of this fit function is the ensured, correct long-term limit of one. The function has been finally applied as template for the adaptation function a(λ,n) of the SMART method (see Chapters 2.4 and 2.5). The fit function was optimized for the range n≤20. If an extended monitoring window of the SMART method is intended (i.e., significantly higher than n_max_=20), one might re-optimize the adaptation function a(λ,n) by refitting Eqs. (10) and (12) with a variable exponent and over the extended range of n. Although, the best fit to an extended MDCI-95% function (n=50) remains very similar: 1+0.986·n**^-0.452^** with an R**^2^**=0.9981. (In the case of MDCI-99%, the best fit is 1+1.5855·n**^-0.479^** with R**^2^**=0.9997). The convincing consensus among both mathematical approaches and the well-matching fit function provide a very reliable basis for the presented SMART method.

The second conclusion regards to the remaining distinct amount of statistical uncertainty in the determination of root mean square metrics based on sample sizes up to about n=50. This uncertainty needs to be considered in limit definitions applied to limited retrospective IQC data. At n=2 the uncertainty (or error) in RMSTD determination can reach about 100% of the true RMSTD value that is exactly obtainable by very large n only. The (hypothetical) large-scale limit L_Δ_ can be chosen close to this true RMSTD value. The findings also confirm the assumption that the upper limits of RMSD/RMSTD results must increase at fewer n and finally culminate in the limit for single control measures L_SMC_ at n=1. According to Chapter 2.1, the ratio L_SMC_ / L_Δ_ of the maximally diverging limits is defined in this study by λ, which lies in the range 1.8-2.5. The fit function F(n) and the equivalent adaptation function a(λ,n) of SMART have been utilized to draw the course of optimal intermediate limits between L_SMC_ and L_Δ_.

The curvature of a(λ,n) or F(n) directly provides the relative amount of the statistical uncertainty at any n by about 

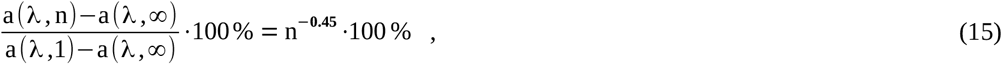

where a(λ,∞)=1. Thus, 30% of the decrease of the adaptation function a(λ,n) from λ (at n=1) towards 1 (for n→∞) still remains at n=15. In other words, the effective range between the maximally diverging applied SMART limits (for n=1 resp. n=15) is just 70% of the entire value of λ. Nevertheless, a monitoring window of n=15 seems to be an optimal trade-off to ensure a fast response to suspicious IQC states. If a larger monitoring window of SMART is preferred, the remaining statistical uncertainties at n=20 and n=30 would be 26% and 21.6%, respectively.

### 3.2 Final remarks on the SMART method

The novel SMART method is especially intended to monitor a limited available number of control measures. However, the monitored window of most recent control measures should not be significantly less than 15 values. During the initial phase, SMART can nevertheless be applied to a smaller data set without modification. The method is most efficient utilizing control samples with known target values. However, a feasible target value could also be self-determined during a pre-analytical evaluation period.

Although the underlying mathematical approaches (Chapter 2.3) may appear complex, the SMART method alone is straightforward. At each application, the SMART method evaluates up to 8 limit tests of the RMSTD values of retrospective SMC charts with increasing considered numbers of recent control measures. The applied limits of the RMSTD results become more and more strict, facilitated by the increasing statistical significance of longer sample series. The limits converge from the tolerant L_SMC_ towards the stricter L_Δ_. Thus, SMART can even quickly respond to suspicious shifts or trends of recent SMC values that are still within the ±L_SMC_ limit around the target value.

One of the main advantages of SMART emerges from the feature that only one prescribed maximum permissible limit per analyte is sufficient to realize the entire IQC - including a fast detection of critical baseline trends as well as single critical outliers. This prescribed limit is either L_SMC_ or L_Δ_, whereas the undefined co-limit is implicitly considered by λ. Hence, the only other necessary parameter is λ, which lies within the range 1.8-2.5 for (almost) all present analytes in clinical chemistry (see Chapter 2.1). Please mention that a suitable choice of λ depends on the preferred variant of SMART. A value of λ near the lower boundary (λ=1.8) is recommended for SMART utilizing L_SMC_ and near the upper boundary (λ=2.5) for SMART utilizing L_Δ_, respectively. The design of the method provides a nuanced interpretation of outliers by alert levels. Thus, the applied prescribed limit for an analyte can be chosen rather strict. It is, however, reasonable to grant a small addition (“bonus”) to further account for some tolerance in the IQC procedure (i.e., permissible heteroscedasticity and baseline drift). The Sigma Metric [13] of the particular measuring procedure may facilitate the decision on the quantity of this addition.

As explained in Chapter 2.2, the recommended number of limit tests is 8 for a monitoring window of n=15 recent SMC values. This SMART plan would simultaneously evaluate 8 retrospective sample series with sizes of n={1,3,5,7,9,11,13,15} (applied after each new SMC measure). In the case of just 6 tests, recommended sample sizes of RMSTD tests would be n={1,3,6,9,12,15} accordingly. In principle, a default number between 6 and 8 limit tests appears optimal. Test frequencies outside this range may require modifications of the definitions of alert levels given in the next chapter. SMART plans, which utilize a retrospective monitoring window of more than 20 SMC values and at least 8 limit tests, should benefit from the optional scoring scheme introduced in Chapter 3.4.

SMART is highly recommended as the new default IQC procedure of the German Rili-BAEK. A change would solve all of the distinct issues of the present mandatory IQC version as discussed in [16]. Further, the current concept of an RA, which is solely applied at the ends of consecutive data-collection periods of 1-3 months, would become obsolete. It would be replaced by the sliding monitoring window of SMART.

### 3.3 The alert levels of SMART

The basic idea behind the definition of alert levels is that a moderate outlier usually breaks just one limit, whereas one distinct outlier breaks multiple SMART limits at once. Particularly, the most recent SMC value will be considered in each of the applied limit tests; hence, it could solely cause multiple limit overruns during one call of SMART. The extended monitoring of SMC data by multiple limit tests thus allows a nuanced interpretation of outliers. In fact, sparse and moderate outliers (exceeding 1-2 test limits) are indeed “welcome” as early problem indicators, and the SMART limits can be defined rather strict.

Moreover, SMART can detect and evaluate a sequence of recent SMC measures that are still below the L_SMC_ limit - but finally fail to meet one or more of the stricter subsequent limits. Nevertheless, the first limit test (L_SMC_), just applied to the current measure, is generally most important, and it has to be more prominently assessed than all other limit tests.

At the end of each call of SMART, the results of all limit tests are processed to reveal the present alert state of the IQC. These alert states range from pre-failure warnings to critical alerts. Table 1 provides a suitable maximum number of 7 distinguishable state levels for an IQC utilizing just one control material. These gradual levels can be used to specifically define preventive actions and detailed instructions for operators.

If two control samples are used and analyzed simultaneously, even more robust IQC state evaluations of the monitored system are possible. Table 2 provides level definitions and suggested instructions for an IQC with two control samples and 6-8 limit tests.

**Table 2:** Level definitions and suggested instructions for an IQC with two simultaneously used control samples (CS)

| Level | Condition | Suggested instructions |
| --- | --- | --- |
| 0 | No violations of limits | Proceed |
| 1 | Only one CS with an outlier at $n=1$ and no other violations (single warning) | Visual inspections of devices, utilities, and CS |
| 1 | Up to two limit violations except at $n=1$ of one or both CS (trend warning) | Exchange of CS and probably other expendable items; extended backup storage of measured patient samples (if reasonable) |
| 2 | Only one CS with an outlier at $n=1$ and up to one other violation in each CS | Educated visual and logged inspections; new CS aliquot; repeat of SMC; further actions might be analyt-specific according to medical relevance, etc. |
| 3 | Both CS with an outlier at $n=1$ and up to one further violation in each CS | Educated visual and logged inspections; cleaning; recalibration; new CS aliquots; repeat of both SMC |
| 4 | Both CS with more than two limit violations each; but no outliers at $n=1$ | Detailed inspection including dis-/reassembly and cleaning of units; exchange of CS and reagents; replacement of exchangeable items or preventive maintenance; recalibration and repeat of both SMC; (inspection of all utility devices; contact lot manufacturer) |
| 5 | An outlier at $n=1$ in combination with at least two further limit violations of one or both CS | Detailed inspection including dis-/reassembly and cleaning of units; exchange of CS and reagents; replacement of exchangeable units or preventive maintenance; recalibration and repeat of both SMC; (inspection of all utility devices; contact lot manufacturer) |
| 6 | Exceeding a predefined maximum number of level-4 or level-5 events within, e.g., 100 days of operation | Reportable incident; re-evaluation of all performance metrics of the device/method |

If an alert level of 4 or higher was reached (and the causing problem has been solved), a restart of SMART by rejecting of all previous SMC data becomes necessary. During the new initial phase one might treat the missing retrospective SMC values within the monitoring window (default n_max_=15) as being equal to the target value. To ensure an efficient IQC even at the very beginning, it is highly recommended to start SMART with a predefined series of n_max_-1 repeats of a better suited value y_dummy_ instead. An adequate value of y_dummy_ is simply the sum of the target value and the strictest limit, which is L_Δ_, thus y_dummy_=y_0_+L_Δ_ or y_dummy_=y_0_+L_SMC_/λ.

How to deal with outliers? Outliers are important state indicators and statistical elements. They should never be ignored even if the SMC measure was directly repeated. Anyway, if an outlier has clearly been dedicated to an apparent exceptional mistake or a compromised control sample; this particular outlier should be rejected. Further, in response to an outlier, the number of permitted direct retries of an SMC has to be strictly limited to avoid repeated measures until a putatively acceptable result is achieved. If nothing has really been changed on the system (except cleaning or recalibration), just one direct repeat of an SMC is maximally permissible.

The optimal number of limit tests, the alert levels, and the suggested instructions were defined by theoretical consideration. Own practical experience during long-term application may lead to further optimizations of these settings.

### 3.4 An optional score for extended monitoring windows

In cases of monitoring windows considering more than about 20 most recent SMC values (N=n_max_, N>20), it might become beneficial to additionally account for the recentness of a failed limit test. This can be achieved by a weighting of limit tests in terms of topicality of the evaluated data. A suitable weight w_n_ is equal to the inverse square root of n, which is the sample size of a particular RMSTD-limit test (with considered retrospective SMC values 1,…,n). The number of limit tests per application of SMART should be at least 8.

The weights of failed limit tests yield a score. The score consists of two numbers due to the extraordinarily prominent single-value limit test for n=1. The first (S_n=1_) is an integer, which defaults to zero but increases by 1 for a failed test at n=1. The second score (S_n>1_) is the normalized sum of the weights of all failed limit tests (except the first one at n=1): 

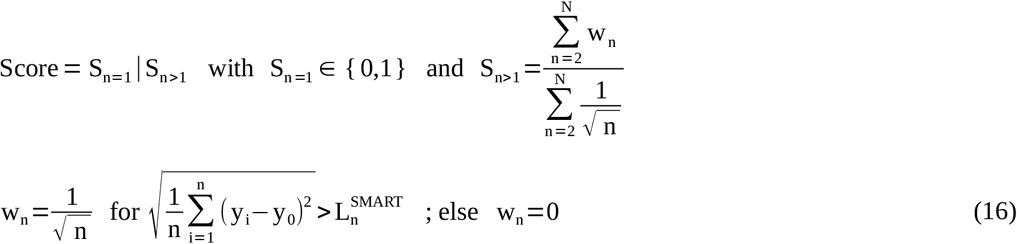

The score is also applicable and recommended for the parallel use of two control samples. Here, the first score can reach two. Regarding S_n>1_, the weights of each control sample are summed up separately; though, the weights of one control sample are considered to be negative (S_n>1_ with minus sign). Table 3 provides suggested definitions of alert levels based on the scoring scheme for one of the smallest SMART plans suitable for scoring.

**Table 3:** Definitions of alert levels for two concurrently used control samples utilizing the optional scoring scheme. The dedicated SMART plan is: n_max_=21 with 8 limit tests at n={1,3,6,9,12,15,18,21}.

| Level | S(n=1) | S(n>1) |
| --- | --- | --- |
| 0 | 0 | 0 |
| 1 | 1 | 0 |
| 1 | 0 | [-0.3,0.3] |
| 2 | 1 | [-0.24,0.24] |
| 3 | 2 | [-0.24,0.24] |
| 4 | 0 | <-0.3 or >0.3 |
| 5 | 1 or 2 | <-0.24 or >0.24 |
| 6 | see Table 2 | see Table 2 |

## 4 Conclusion

The study quantitatively revealed that a significant statistical uncertainty in the determination of RMSTD or RMSD metrics must be considered, if the sample size (n) is below about 50. Thus, applied statistical IQC limits need to be adapted according to the size of monitored data. A transition curve of optimal IQC limits is given, which connects the maximum permissible limit for single measures (n=1) with the RMSTD limit yielded by large-scale statistics (n→∞).

The findings led to the development of the novel SMART method intended for small monitoring windows. The method manages the entire IQC with only one mandatory limit statement per analyte. SMART can distinguish between up to 7 alert levels due to multiple limit tests. If a retrospective monitoring window of more than 20 values is preferred, an optional weighting of failed single tests can be applied to further improve the topicality of potential critical alert levels.

## Data Availability

All data sources are mentioned in the reference list.

## Abbreviations

CI: confidence interval
CI_Δ_^up^: upper limit of the confidence interval of Δ
Δ: the “true” RMSTD value determined using large amounts of data
GPD: Gaussian probability density
IQC: internal quality control
L_SMC_: IQC limit applied to single measures of a control material
L_Δ_: IQC limit dedicated to the true (long-term) RMSTD
MDCI: multidimensional confidence interval
RA: retrospective (statistical) analysis of SMC data
Rili-BAEK: Guideline of the German Medical Association on Quality Assurance in Medical Laboratory Examinations
RMSD: root mean square deviation (empirical standard deviation)
RMSTD: root mean square total (or target) deviation with respect to a known target value
SMART: Statistical Monitoring by Adaptive RMSTD Tests
SMC: single measurement of a control sample

## Research Funding

This research did not receive any specific grant from funding agencies in the public, commercial, or not-for-profit sectors.

